# Lumipulse G SARS-CoV-2 Ag Assay Evaluation for SARS-CoV-2 Antigen Detection Using 594 Nasopharyngeal Swab Samples from Different Testing Groups

**DOI:** 10.1101/2021.01.26.21250533

**Authors:** Giulia Menchinelli, Licia Bordi, Flora Marzia Liotti, Ivana Palucci, Maria Rosaria Capobianchi, Giuseppe Sberna, Eleonora Lalle, Lucio Romano, Giulia De Angelis, Simona Marchetti, Maurizio Sanguinetti, Paola Cattani, Brunella Posteraro

## Abstract

Compared to RT-PCR, lower performance of antigen detection assays, including the Lumipulse G SARS-CoV-2 Ag assay, may depend on specific testing scenarios. We tested 594 nasopharyngeal swab samples from individuals with COVID-19 (RT-PCR cycle threshold [Ct] values ≤40) or non-COVID-19 (Ct values ≤40) diagnoses. RT-PCR positive samples were assigned to diagnostic, screening, or monitoring groups of testing. With a limit of detection of 1.2 × 10^4^ SARS-CoV-2 RNA copies/ml, Lumipulse showed positive percent agreement (PPA) of 79.9% (155/194) and negative percent agreement of 99.3% (397/400), whereas PPAs were 100% for samples with Ct values of <18 or 18–<25 and 92.5% for samples with Ct values of 25–<30. By three groups, Lumipulse showed PPA of 87.0% (60/69), 81.1% (43/53), or 72.2% (52/72), respectively, whereas PPA was 100% for samples with Ct values of <18 or 18–<25, and was 94.4%, 80.0%, or 100% for samples with Ct values of 25–<30, respectively. RT-PCR positive samples were also tested for SARS-CoV-2 subgenomic RNA and, by three groups, testing showed that PPA was 63.8% (44/69), 62.3% (33/53), or 33.3% (24/72), respectively. PPAs dropped to 55.6%, 20.0%, or 41.7% for samples with Ct values of 25–<30, respectively. All 101 samples with a subgenomic RNA positive result had a Lumipulse assay’s antigen positive result, whereas only 54 (58.1%) of remaining 93 samples had a Lumipulse assay’s antigen positive result. In conclusion, Lumipulse assay was highly sensitive in samples with low RT-PCR Ct values, implying repeated testing to reduce consequences of false-negative results.

## INTRODUCTION

Antigen testing has recently been added to the landscape of clinical laboratory methods to detect and combat the severe acute respiratory syndrome coronavirus 2 (SARS-CoV-2), which is the notorious cause of coronavirus disease 2019 (COVID-19) (https://www.cdc.gov/coronavirus/2019-ncov/lab/resources/antigen-tests-guidelines.html#anchor_1597523027400). Like the molecular— relying on real-time reverse-transcription polymerase chain reaction (RT-PCR) and, to date, the standard method for the etiological COVID-19 diagnosis—antigen testing detects the presence of SARS-CoV-2 in the acute infection phase only (1). Unlike the molecular or antigen, antibody testing is relevant in the convalescent or recovered infection phases only (1).

Theoretically, antigen-based assays are advantageous in terms of fast turnaround times and reduced costs but are less sensitive than RT-PCR-based assays (2). Additionally, the former have the disadvantage to provide false-positive results, which leads false-positive patients to be managed as patients with true SARS-CoV-2 infection (3, 4). However, the false-positive result likelihood seems to depend on specific testing scenarios (e.g., those to identify infected persons who are asymptomatic and without known or suspected exposure to SARS-CoV-2) (https://www.cdc.gov/coronavirus/2019-ncov/lab/resources/antigen-tests-guidelines.html#anchor_1597523027400). To mitigate this issue, the European Centre for Disease Prevention and Control (ECDC) recommends antigen-based assays to be not only carefully selected but also validated before their implementation in clinical practice (5). Since June 2020, the Lumipulse G SARS-CoV-2 Ag (Fujirebio, Tokyo, Japan), detecting SARS-CoV-2 nucleocapsid (N) protein, is being used in Japan where, according to the Japanese Ministry of Health, Labour and Welfare policy (4), a positive antigen test result is enough to definitively diagnose COVID-19 without PCR—which is instead mandatory in European countries to confirm positive antigen results (5). Two independent studies by Hirotsu et al. (6, 7) reported on the performance of the Lumipulse G SARS-CoV-2 Ag assay (hereafter referred as the Lumipulse assay) using nasopharyngeal swab samples. Serial or individual samples from 11 RT-PCR positive (SARS-CoV-2 infected) patients and 215 RT-PCR negative (SARS-CoV-2 uninfected) patients (6) or 27 serial samples from one patient with persistent SARS-CoV-2 RNA shedding during hospitalization (7) were used. In both studies, samples with high viral load (corresponding to low values of RT-PCR cycle threshold [Ct]—an accredited measure of virus [5]) or samples collected in the early infection phase showed complete concordance between Lumipulse and RT-PCR results.

With the aim to fully understanding its usefulness, we evaluated the Lumipulse assay with 594 individuals’ nasopharyngeal swab samples assigned to different testing groups (i.e., including early or late infection patients). To this end, we compared Lumipulse assay antigen results with those of RT-PCR assay targeting SARS-CoV-2 genomic RNA (usually used as an indicator of viral presence). In parallel, RT-PCR positive samples were analyzed for the presence of subgenomic RNA (recently proposed as an indicator of active viral replication) to support Lumipulse assay’ results.

## MATERIALS AND METHODS

### Study design and clinical samples

This study was conducted at the Fondazione Policlinico Universitario A. Gemelli IRCCS (FPG) and was approved by the FPG Ethics Committee (reference number 49978/20). Informed consent was obtained from all participants before including their samples in the study. We included nasopharyngeal swab samples from patients/individuals (≥18-year aged) presenting at and/or admitted to our institution during a 2-week period in December 2020. Samples were from laboratory-confirmed COVID-19 (*n* = 194) or non-COVID-19 (*n* = 400) diagnoses, which relied, respectively, on positive (Ct values of ≤40) or negative (Ct values of >40) results obtained using the Seegene Allplex 2019-nCoV, the DiaSorin Simplexa COVID-19 Direct, or the Roche Diagnostics Cobas SARS-CoV-2 Test RT-PCR assays (8–10). For example, the Seegene Allplex 2019-nCoV assay is a single-tube assay targeting the envelope (E), RdRP (RNA-dependent RNA polymerase), and N SARS-CoV-2 genes and running on a Bio-Rad CFX96 Real-time Detection system. Based on Ct values—i.e., numbers of cycles the fluorescent signal crosses the threshold for positive detections—the Seegene software automatically analyzes RT-PCR results. By this assay, a Ct value ≤40 for at least one of two viral genes (i.e., RdRP and N) or for the E gene alone indicates, respectively, the certain or presumptive presence of SARS-CoV-2 RNA in the sample. No positive samples only for E gene were included in the study. In view of relatively lower performance of the DiaSorin Simplexa COVID-19 Direct assay (8, 11), samples (*n* = 39) initially tested with this assay were retested with the Seegene Allplex 2019-nCoV assay to confirm (positive) results. Likewise, samples with discordant results between the RT-PCR and the Lumipulse assays (see below) were confirmed as positive (*n* = 39) or negative (*n* = 3) by retesting as previously described (12).

For stratification purposes (5), we selected positive samples based on their Ct values (i.e., 11.2– 39.9) to include samples with different viral load levels. These samples were characterized for the inclusion in three testing groups, namely diagnostic, screening, and monitoring groups, which were in substantial accordance with the definitions reported in the interim technical guidance by the Centers for Disease Control and Prevention (CDC) for rapid SARS-CoV-2 antigen testing (https://www.cdc.gov/coronavirus/2019-ncov/lab/resources/antigen-tests-guidelines.html#anchor_1597523027400). Accordingly, diagnostic or monitoring groups included persons who had signs or symptoms (i.e., clinical illness) consistent with COVID-19, who had no clinical illness but a recent known or suspected exposure to SARS-CoV-2, or who had a previous laboratory-confirmed COVID-19 diagnosis, whereas the screening group included persons who were asymptomatic and without known or suspected exposure to SARS-CoV-2. All positive samples were further stratified in five groups based on RT-PCR Ct values (<18, 18–<25, 25–<30, 30–<35, and 35–40).

All samples originally collected into universal transport medium (UTM; Copan, Brescia, Italy) were portioned in aliquots that were kept at 4°C until testing with the Lumipulse assay (see below), which was always performed within 2–4 hours from the time samples were subjected to RT-PCR for detecting SARS-CoV-2 genomic RNA as above described. In parallel, additional aliquots from the same samples were frozen at −80°C until testing for SARS-CoV-2 subgenomic RNA (see below). Furthermore, we used archived frozen samples (RT-PCR negative) as a matrix to generate contrived samples for the Lumipulse assay’s analytical sensitivity determination (see below).

### Lumipulse assay for SARS-CoV-2 antigen detection

The Lumipulse assay quantitatively detects SARS-CoV-2 N protein in clinical samples (e.g., nasopharyngeal swab) by a specific two-reaction chemiluminescence-based immunoassay method on the Lumipulse G1200 automated immunoassay analyzer (Fujirebio). In the first reaction, the sample (or the SARS-CoV-2 Ag calibrator) and the sample treatment solution are added to an anti-SARS-CoV-2 monoclonal antibody-coated magnetic particle solution, and then incubated for 10 min at 37°C to allow formation of specific antigen-antibody immunocomplexes. In the second reaction (accessed after washing), an alkaline phosphatase-labelled anti-SARS-CoV-2 monoclonal antibody solution is added and incubated for 10 min at 37°C to allow specific binding to the antigen of aforementioned immunocomplexes, and then to form additional immunocomplexes. Finally (after washing), a substrate solution is added and incubated for 5 min at 37°C, and the resulting chemiluminescence signals are automatically read by the analyzer and used to calculate the SARS-CoV-2 antigen’s amount in the sample through the interpolation with a SARS-CoV-2 Ag calibrator curve.

We determined the limit of detection (LOD) of the Lumipulse assay according to a previously described protocol (9). Briefly, aforementioned contrived samples were spiked with a dilution series of Vero E6 cell-cultured SARS-CoV-2 (INMI-1 strain) at a concentration range of 1.0 × 10^5^ 50% tissue culture infective dose (TCID_50_)/ml (4.0 × 10^8^ RNA copies/ml) to 1.0 TCID_50_/ml (4.0 × 10^3^ RNA copies/ml), and then tested in replicates (Fig. S1). For each sample, SARS-CoV-2 RNA was amplified by RT-PCR in Rotor-GeneQ Real-Time cycler (Qiagen, Hilden, Germany), using the RealStar SARS-CoV-2 RT-PCR kit 1.0 (Altona Diagnostic GmbH, Hamburg, Germany). RNA copies/ml were calculated through a standard curve prepared with serially diluted EURM-019 single-strand SARS-CoV-2 RNA fragments (https://crm.jrc.ec.europa.eu/p/EURM-019). Thus, we plotted the probability (y-axis) against the SARS-CoV-2 concentration’s logarithm (x-axis), and we calculated the 95% LOD value, which was the lowest concentration at which the replicates yielded positive detection 95% of the time (Fig. S1).

Before testing with the Lumipulse assay, samples were centrifuged at 3000 × g for 15 min to allow separation of the supernatant from the remaining viscous UTM material, and 100 µl were analyzed for the antigen quantification as above described. Samples with an antigen level exceeding the detection limit (i.e., 5000 pg/ml) were diluted, and dilutions were used to quantify the original samples’ antigen levels based on the dilution factor. Results were interpreted using a cutoff of 1.34 pg/ml as established by the Lumipulse assay’s manufacturer, and were expressed as negative (<1.34 pg/ml), gray-zone positive (1.34–10 pg/ml), or positive (>10–>5000) results, respectively. For convenience reasons, antigen concentrations >5000 pg/ml were rounded to 5000 pg/ml.

### RT-PCR assay for SARS-CoV-2 subgenomic RNA detection

To determine the presence of SARS-CoV-2 subgenomic RNA (i.e., E gene subgenomic RNA), samples were subjected to a previously developed in-house RT-PCR assay (13). This is an adaptation from the method described by Wölfel et al. (14) that looks specifically at the E gene subgenomic RNA to indicate active virus infection/transcription (15). Briefly, SARS-CoV-2 RNA (also including genomic RNA) was extracted from samples using the Seegene Nimbus automated system and then used for the RT-PCR assay. This was performed with the Qiagen OneStep RT-PCR kit (Qiagen, Valencia, CA) and a 25-μl reaction volume containing 600 nM concentration each of primers (sgE_SARS-CoV2_F 5’-CGATCTCTTGTAGATCTGTTCTC-3’; sgE_SARS-CoV2_R 5’-ATATTGCAGCAGTACGCACACA-3’) and 200 nM concentration of probe (sgE_SARS-CoV2_P 5’-FAM-ACACTAGCCATCCTTACTGCGCTTCG-BBQ-3’). Thermal cycling consisted of 30 min at 50°C for reverse transcription, followed by 15 min at 95°C and subsequent 45 cycles each of 10 s at 95°C, 15 s at 55°C, and 30 s at 72°C.

### Data collection and analysis

Data were presented as numbers with percentages or as means ± standard deviation (SD), as appropriate. To determine Lumipulse assay’s LOD, the MedCalc statistical software (MedCalc Software Ltd, Ostend, Belgium) was used to convert RT-PCR positive detection proportion into a “probability unit” (or “probit”). Lumipulse assay’s results were categorized as positive, gray-zone positive, or negative and, then, compared using a one-way analysis of variance (ANOVA) with the Tukey’s multiple-comparison test. For Lumipulse assay’s or subgenomic RNA assay’s results, differences between *a priori* established groups were assessed using the chi-square test or the Student’s *t*-test, as appropriate. Percent agreement values, with their respective confidence intervals (CIs), were calculated comparing Lumipulse assay’s or subgenomic RNA assay’s results with those obtained by the reference method (i.e., genomic RNA RT-PCR assay). Correlation between antigen levels (as determined by the Lumipulse assay) and Ct values (as determined by the reference method) was assessed using the Spearman’s correlation coefficient. Statistical analysis was conducted using Stata 15 (StataCorp, College Station, TX) or GraphPad Prism 7 (GraphPad Software, San Diego, CA) software. *P* <0.05 was considered statistically significant.

## RESULTS

### SARS-CoV-2 antigen (Lumipulse assay) versus genomic or subgenomic RNA (RT-PCR assay) testing

First, we determined the analytical capability of the Lumipulse assay, a recently marketed assay for SARS-CoV-2 N protein detection in European countries. As shown in Fig. S1, the LOD was 2.95 TCID_50_/ml, corresponding to 1.2 × 10^4^ SARS-CoV-2 RNA copies/ml, at 95% detection probability. Then, 594 nasopharyngeal swab samples, including RT-PCR positive (*n* = 194) or negative (*n* = 400) samples, were tested with the Lumipulse assay.

#### i) Overall performance

Using SARS-CoV-2 RNA genomic RT-PCR assay as the reference method (Table 1), the Lumipulse assay detected 155 of 194 samples as positive (antigen concentration, ≥1.34 pg/ml) and 397 of 400 samples as negative (antigen concentration, <1.34 pg/ml). This resulted in a positive percent (PPA) of 79.9% (95% confidence interval (CI), 73.6– 85.3) and a negative percent agreement (NPA) of 99.3% (95% CI, 97.8–99.8), respectively. Of 155 samples, 29 (18.7%) were positive within the gray-zone (antigen concentration, 1.34–10 pg/ml), which defines an antigen positivity extent necessitating to be confirmed by RT-PCR. As depicted in Fig. 1 and detailed in Table 1, we stratified Lumipulse assay’s results according to RT-PCR Ct values. Thus, we found significant differences in the mean Ct value ± SD for 126 samples with antigen-positive results (21.95 ± 6.03) as compared to 29 samples with antigen (gray-zone)-positive results (30.85 ± 3.19) and to 39 samples with antigen-negative results (33.79 ± 2.39), respectively (ANOVA with Tukey’s multiple-comparison test; *P* <0.0001 for both comparisons) (Fig. 1). Interestingly, PPAs between Lumipulse assay’s and RT-PCR assay’s results were 100% for samples with Ct values of <18 (*n* = 38) or 18–<25 (*n* = 49) and 92.5% for samples with Ct values of 25–<30 (*n* = 37). For 31 of 155 samples with Ct values of 30–<35 (*n* = 23) or 35–40 (*n* = 8), PPAs dropped to 47.9% and 42.1%, respectively. More interestingly, 24 (82.8%) of 29 antigen (gray zone)-positive results regarded samples with Ct values ranging from 25 to 35, whereas three (100%) of three antigen (gray zone)-positive results regarded (antigen false-positive) samples with Ct values >40.

**TABLE 1.** Comparison of Lumipulse assay results with the RT-PCR assay results stratified by Ct values for SARS-CoV-2 detection

| RT-PCR assay<sup>a</sup> | Lumipulse assay<sup>b</sup> |  |
| --- | --- | --- |
| Ct values (no. of results) | No. of positive (including gray-zone positive) results | Percent agreement (95% confidence interval) |
| ≤40 (194) | 155 (29) | 79.9 (73.6–85.3) |
| <18 (38) | 38 (0) | 100.0 (90.7–100.0) |
| 18–<25 (49) | 49 (1) | 100.0 (92.7–100.0) |
| 25–<30 (40) | 37 (10) | 92.5 (79.6–98.4) |
| 30–<35 (48) | 23 (14) | 47.9 (33.3–62.8) |
| 35–40 (19) | 8 (4) | 42.1 (20.3–66.5) |
| >40 (400) | 3 (3) | 99.3 (97.8–99.8) |

Abbreviations: RT-PCR, real-time reverse-transcription polymerase chain reaction; SARS-CoV-2, severe acute respiratory syndrome coronavirus 2; Ct, cycle threshold.
<sup>a</sup> SARS-CoV-2 RT-PCR testing was performed on 594 individuals' nasopharyngeal swab samples. A positive result (i.e., a Ct ≤40) for at least one of two viral targets with the Seegene Allplex 2019-nCoV (nucleocapsid [N] and RdRP [RNA-dependent RNA polymerase]), the DiaSorin Simplexa COVID-19 Direct (S [spike] and ORF1ab [open reading frame 1ab]), or the Roche Diagnostics Cobas SARS-CoV-2 Test (E [envelope] and ORF1a) assays indicated the presence of SARS-CoV-2 RNA in individual's nasopharyngeal swab samples (*n* = 194). Samples with a Ct >40 for the mentioned genes were considered negative (*n* = 400).
<sup>b</sup> SARS-CoV-2 antigen testing was performed on 594 individuals' nasopharyngeal swab samples with the Lumipulse G SARS-CoV-2 Ag assay. Using the manufacturer's cutoff of 1.34 pg/ml, results were interpreted as gray-zone positive or positive when antigen concentrations were 1.34–10 and >10–5000 pg/ml, respectively.

**FIG 1.**
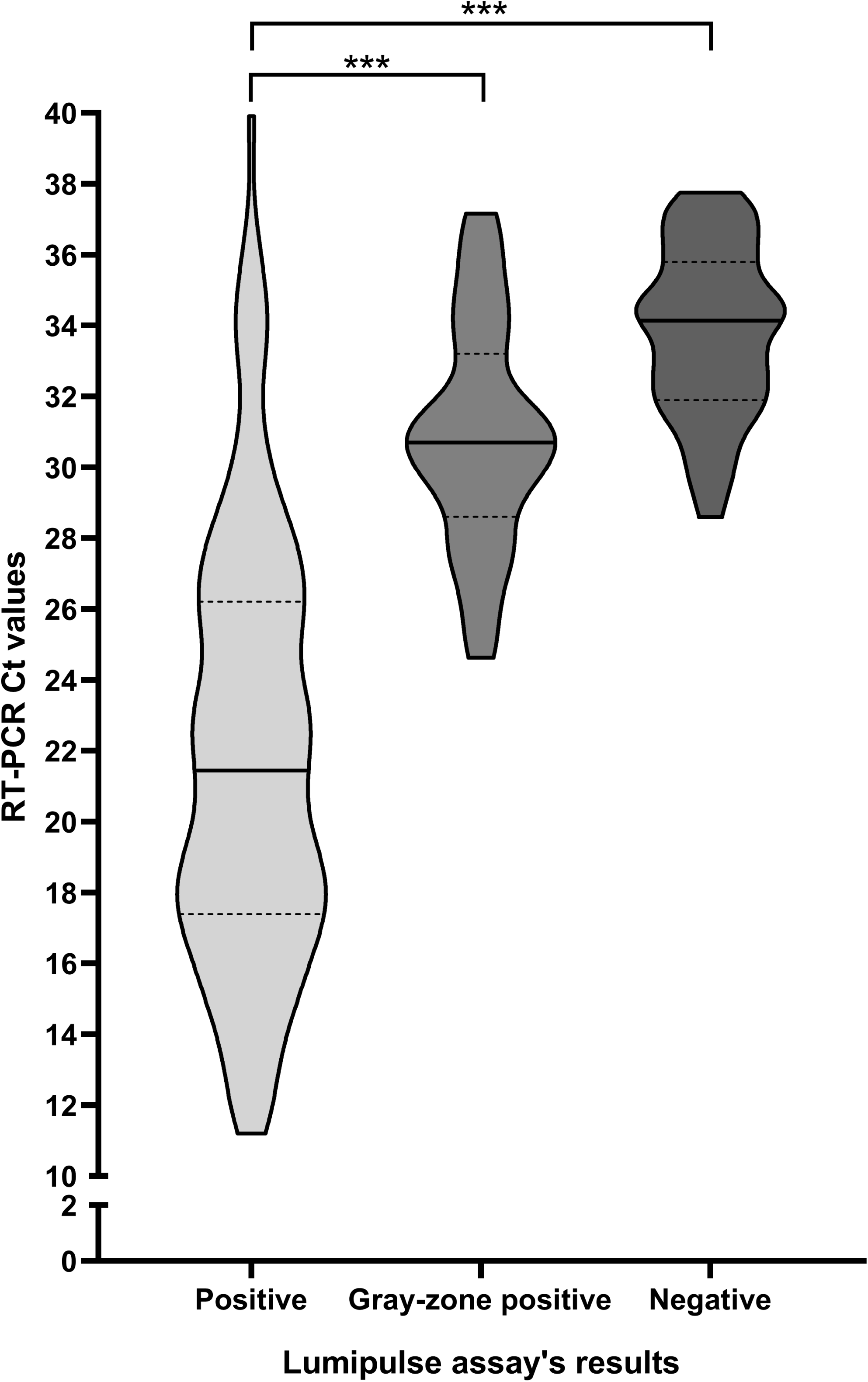
Distribution of Lumipulse assay’s results according to RT-PCR assay’s Ct values in 194 nasopharyngeal samples. Results are presented as positive (>10–5000), gray-zone positive (1.34–10 pg/ml), or negative (<1.34 pg/ml), respectively. In each violin plot, solid line indicates the mean Ct value (21.95, 30.85, and 33.79, respectively) and the area between dotted lines indicates the standard deviation value (6.03, 3.19, and 2.39, respectively). Asterisks indicate statistically significance (*P* <0.0001) between result groups, as established using a one-way analysis of variance (ANOVA) with the Tukey’s multiple-comparison test.

#### ii) Performance by different testing groups

Table 2 shows the results of 194 antigen-positive samples—overall described in Table 1—stratified by the diagnostic (*n* = 69), screening (*n* = 53), or monitoring (*n* = 72) groups of testing for 194 patients with laboratory-confirmed COVID-19 diagnosis. Only for the monitoring group, samples used in the study were not the same as those at the COVID-19 diagnosis time; thus, this group included COVID-19 patients who were tested during the course of disease. Conversely, 122 patients in the two remaining groups were tested at early disease phases. Table 2 also shows the results from SARS-CoV-2 subgenomic RNA detection that was performed in parallel on the 194 samples.

Regarding antigen detection results, PPA with the reference method (i.e., SARS-CoV-2 RNA genomic RT-PCR assay) was 87.0% (95% CI, 76.7–93.9; 60/69 results), 81.1% (95% CI, 68.0– 90.6; 43/53 results), or 72.2% (95% CI, 60.4–82.1; 52/72 results) in diagnostic, screening, and monitoring groups, respectively. Consistent with that shown in Table 1, PPA was 100% for samples with Ct values of <18 or 18–<25 in all three testing groups, and was 94.4%, 80.0%, or 100% for samples with Ct values of 25–<30 in diagnostic, screening, and monitoring groups, respectively. Regarding subgenomic RNA detection results, PPA with the reference method was 63.8% (95% CI, 51.3–75.0; 44/69 results), 62.3% (95% CI, 47.9–75.2; 33/53 results), or 33.3% (95% CI, 22.7–45.4; 24/72 results) in diagnostic, screening, and monitoring groups, respectively. Unlike antigen detection results, PPAs for samples with Ct values of <18 or 18–<25 were, respectively, 86.7% and 100% in the diagnostic group, 100% and 94.1% in the screening group, and 100% and 83.3% in the monitoring group. Interestingly, in all three groups, PPAs dropped to 55.6%, 20.0%, or 41.7% for samples with Ct values of 25–<30, and reached 0% for almost all samples with Ct values of 30–<35 or 35–40, respectively. A chi-square test analysis was conducted to compare PPAs between antigen and subgenomic-RNA detections among the three testing groups, and this analysis revealed significant differences for the samples overall (*P* = 0.002, *P* = 0.03, and *P* <0.001, respectively) or the samples with Ct values ranging from 25–<30 or 35–40 (*P* <0.05 for all comparisons).

**TABLE 2.** Positive detections of Lumipulse antigen and subgenomic RNA compared with those of RT-PCR for SARS-CoV-2 in different testing groups^a^

| Results according to indicated RT-PCR Ct values for: |  |  |  |  |  |
| --- | --- | --- | --- | --- | --- |
| Ct values (no. of results) | Lumipulse antigen detection<sup>b</sup> |  | Subgenomic RNA detection<sup>c</sup> |  | *P* value<sup>d</sup> |
|  | No. of results (including gray-zone results) | Percent agreement (95% confidence interval) | No. of results | Percent agreement (95% confidence interval) |  |
| Diagnostic group |  |  |  |  |  |
| <18 (15) | 15 (0) | 100.0 (78.2–100.0) | 13 | 86.7 (59.5–98.3) | 0.14 |
| 18–<25 (20) | 20 (0) | 100.0 (83.2–100.0) | 20 | 100.0 (83.2–100.0) | NA |
| 25–<30 (18) | 17 (1) | 94.4 (72.7–99.9) | 10 | 55.6 (30.8–78.5) | 0.009 |
| 30–<35 (10) | 6 (5) | 60.0 (26.2–87.8) | 0 | 0.0 (0.0–30.8) | 0.003 |
| 35–40 (6) | 2 (0) | 33.3 (4.3–77.7) | 1 | 16.7 (0.4–64.1) | 0.52 |
| All (69) | 60 (6) | 87.0 (76.7–93.9) | 44 | 63.8 (51.3–75.0) | 0.002 |
| Screening group |  |  |  |  |  |
| <18 (15) | 15 | 100.0 (78.2–100.0) | 15 | 100.0 (78.2–100.0) | NA |
| 18–<25 (17) | 17 | 100.0 (80.5–100.0) | 16 | 94.1 (71.3–99.9) | 0.30 |
| 25–<30 (10) | 8 (5) | 80.0 (44.4–97.5) | 2 | 20.0 (2.5–55.6) | 0.007 |
| 30–<35 (8) | 3 (2) | 38.0 (8.5–75.5) | 0 | 0.0 (0.0–36.9) | 0.05 |
| 35–40 (3) | 0 | 0.0 (0.0–70.6) | 0 | 0.0 (0.0–70.6) | NA |
| All (53) | 43 (7) | 81.1 (68.0–90.6) | 33 | 62.3 (47.9–75.2) | 0.03 |
| Monitoring group |  |  |  |  |  |
| <18 (8) | 8 | 100.0 (63.1–100.0) | 8 | 100.0 (63.1–100.0) | NA |
| 18–<25 (12) | 12 (1) | 100.0 (73.5–100.0) | 10 | 83.3 (51.6–97.9) | 0.13 |
| 25–<30 (12) | 12 (4) | 100.0 (73.5–100.0) | 5 | 41.7 (15.2–72.3) | 0.002 |
| 30–<35 (30) | 14 (7) | 46.7 (28.3–65.7) | 1 | 3.3 (0.1–17.2) | <0.001 |
| 35–40 (10) | 6 (4) | 60.0 (26.2–87.8) | 0 | 0.0 (0.0–30.8) | 0.003 |
| All (72) | 52 (16) | 72.2 (60.4–82.1) | 24 | 33.3 (22.7–45.4) | <0.001 |

Abbreviations: RT-PCR, real-time reverse-transcription polymerase chain reaction; SARS-CoV-2, severe acute respiratory syndrome coronavirus 2; Ct, cycle threshold; NA, not applicable.
<sup>a</sup> Groups were established according to the Centers for Disease Control and Prevention (CDC) definitions for testing settings (https://www.cdc.gov/coronavirus/2019-ncov/lab/resources/antigen-tests-guidelines.html#anchor_1597523027400), and were further stratified by viral load (i.e., Ct values) as indicated.
<sup>b</sup> SARS-CoV-2 antigen was detected in nasopharyngeal swab samples of groups' individuals by the Lumipulse G SARS-CoV-2 Ag assay, which provides a 0.01–5000 pg/ml measurement range. Using the manufacturer's cutoff of 1.34 pg/ml, results were expressed as negative, gray-zone positive, or positive when antigen concentrations in the samples were <1.34, 1.34–10, or >10–5000 pg/ml, respectively. Samples with antigen concentrations above 5000 pg/ml were rounded to 5000 pg/ml for convenience reasons.
<sup>c</sup> SARS-CoV-2 subgenomic RNA was detected in nasopharyngeal swab samples of groups' individuals by in-house RT-PCR assay for the presence of replicative (E gene) RNA (Liotti, 2020c).
<sup>d</sup> For comparisons between percent agreement rates.

#### iii) Correlation between antigen levels and RT-PCR Ct values

To corroborate these findings, we assessed antigen levels in relation with the SARS-CoV-2 viral load expressed as RT-PCR Ct values. A Spearman’s correlation analysis was conducted for all 194 samples that tested positive with the RT-PCR assay, which were analyzed according to aforementioned testing groups (i.e., diagnostic, screening, and monitoring). As shown in Fig. 2, we found a significant (negative) association between antigen levels and Ct values in either diagnostic (Spearman’s *ρ* = −0.82; *P* <0.0001), monitoring (Spearman’s *ρ* = −0.76; *P* <0.0001), or screening (Spearman’s *ρ* = −0.72; *P* <0.0001) groups. As it can see, association was relatively stronger in the diagnostic group and less strong in the screening group.

**FIG 2.**
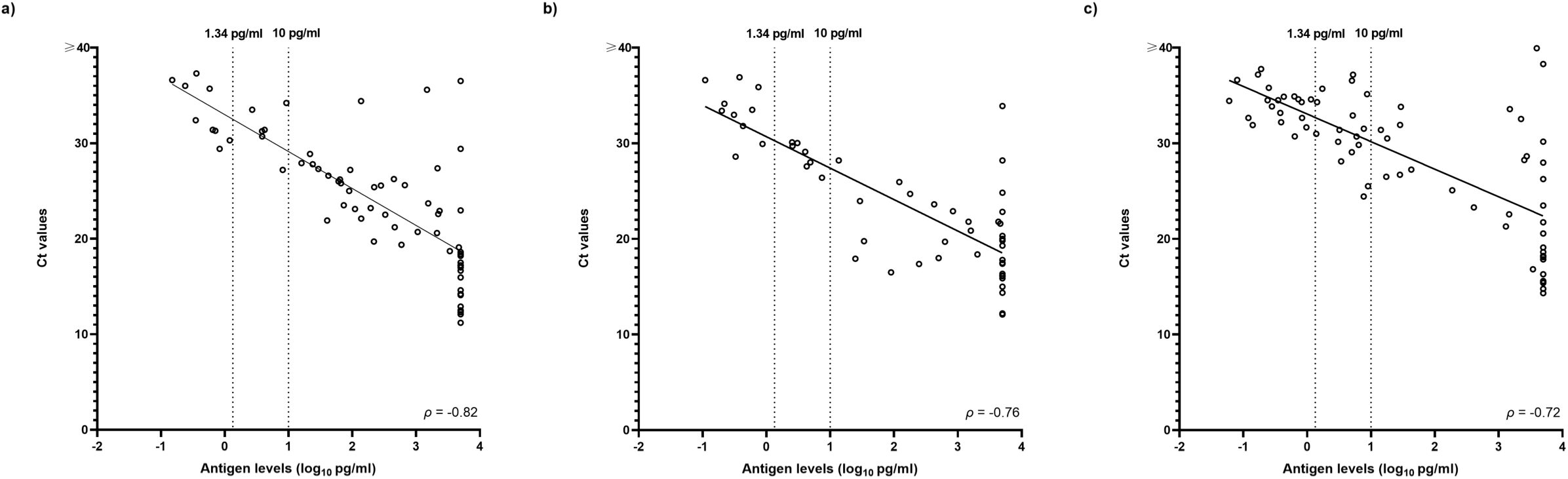
Correlation between the SARS-CoV-2 antigen levels quantified by the Lumipulse assay and the SARS-CoV-2 genomic RNA Ct values obtained with the RT-PCR assay. Analysis was separately conducted for (a) diagnostic, (b) screening, and (c) monitoring testing groups. Antigen concentration is expressed as log_10_ pg/ml. Concentrations of <1.34 pg/ml, 1.34 to 10 pg/ml, and >10 pg/ml were used to interpret Lumipulse assay’s antigen results as negative, gray-zone positive, or positive, respectively.

### Relationship between SARS-CoV-2 antigen and subgenomic RNA

To investigate this issue, we analyzed the characteristics of 194 antigen-positive or -negative samples according to the presence (*n* = 101) or absence (*n* = 93) of subgenomic RNA. As shown in Table 3, all 101 samples with a subgenomic RNA positive result had a Lumipulse assay’s antigen positive result, whereas only 54 (58.1%) of remaining 93 samples had a Lumipulse assay’s antigen positive result. Samples in the subgenomic RNA-positive group had a mean Ct value ± SD—at the RT-PCR assay for genomic RNA—that did not significantly differ from that of samples in the subgenomic RNA-negative group (20.3 ± 4.8 versus 29.9 ± 4.8; Student’s *t*-test; *P =* 0.43). Conversely, the time from COVID-19 diagnosis to testing (mean days ± SD) for samples in the subgenomic RNA-positive group significantly differed from the time for samples in the subgenomic RNA-negative group (1.6 ± 3.3 versus 6.1 ± 7.0; Student’s *t*-test; *P* <0.001).

**TABLE 3.** Characteristics of 194 RT-PCR positive samples that tested positive or negative for the presence of SARS-CoV-2 subgenomic RNA^a^

| Characteristic<sup>b</sup> | Samples with a subgenomic RNA positive result (*n* = 101) grouped as |  | Samples with a subgenomic RNA negative result (*n* = 93) grouped as |  | *P* value<sup>c</sup> |
| --- | --- | --- | --- | --- | --- |
|  | RT-PCR positive/Antigen positive (*n* = 101) | RT-PCR positive/Antigen negative (*n* = 0) | RT-PCR positive/Antigen positive (*n* = 54) | RT-PCR positive/Antigen negative (*n* = 39) |  |
| RT-PCR Ct, mean value ± SD | 20.3 ± 4.8 | NA<sup>d</sup> | 29.9 ± 4.8 | 33.8 ± 2.4 | 0.43 |
| Testing from COVID-19 diagnosis, mean days ± SD | 1.6 ± 3.3 | NA<sup>d</sup> | 6.1 ± 7.0 | 7.7 ± 8.6 | <0.001 |

<sup>a</sup> All samples were from diagnostic (*n* = 59), screening (*n* = 63), or monitoring (*n* = 72) testing groups (see Table 2). Testing for SARS-CoV-2 subgenomic RNA was performed using an in-house RT-PCR assay to assess the presence of replicative (E gene) RNA, as previously described (Liotti, 2020c).
<sup>b</sup> The time period between SARS-CoV-2 RT-PCR (to which Ct values refer) used to diagnose COVID-19 and testing for SARS-CoV-2 subgenomic RNA (and antigen) ranged from 0 days in the diagnostic or screening groups to 32 days in the monitoring group. Only in the last group, consequently, two temporally different samples were tested.
<sup>c</sup> For comparisons between the RT-PCR positive/Antigen positive groups herein listed.
<sup>d</sup> NA, not applicable.

## DISCUSSION

Taking advantage of antigen tests used to detect infection with viruses other than SARS-CoV-2 (16), diagnostics of COVID-19 quickly moved to use non-molecular (i.e., non-RT-PCR) laboratory tests, including the Lumipulse assay (5). Unlike point-of-care antigen test formats (17), Lumipulse assay employs a chemiluminescence-based SARS-CoV-2 antigen quantification (6), which should enhance the test sensitivity to diagnose SARS-CoV-2 infection. To evaluate the Lumipulse assay, we used RT-PCR as the best available comparator method (1), and we showed that the Lumipulse assay had PPA (sensitivity) and NPA (specificity) of ∼80% and 99%, respectively. Consequently, the number of false-positive and-negative results driven by these metrics was 39 (among 194 RT-PCR positive results) and 3 (among 400 RT-PCR negative results), respectively (Table 1). As the Lumipulse assay returns positive results as positive (>10 to 5000 pg/ml) or gray-zone positive (≥1.34 to 10 pg/ml), it is worthy to note that all the three false-positive results fell within the gray-zone range. These results would have been confirmed by RT-PCR if antigen testing had been performed as a frontline diagnostic method at the study time.

In our hands, Lumipulse assay met the minimum performance requirements of ≥80% sensitivity and ≥97% specificity for rapid antigen tests as established by the World Health Organization (https://www.who.int/publications/m/item/covid-19-target-product-profiles-for-priority-diagnostics-to-support-response-to-the-covid-19-pandemic-v.0.1) and, later, agreed by the ECDC (5). Additionally, we showed that the Lumipulse assay’s sensitivity increased from ∼93% to 100% with samples that displayed RT-PCR Ct values below 25–30. As a reflection of high viral load, these values are likely associated with an infectious SARS-CoV-2, contrasting higher Ct values (>30 to 40) that, instead, are likely associated with a non-infectious SARS-CoV-2 (14). A manner for appraising the apparently low performance of the Lumipulse assay as compared to RT-PCR (i.e., ≥90% sensitivity [https://www.who.int/publications/m/item/covid-19-target-product-profiles-for-priority-diagnostics-to-support-response-to-the-covid-19-pandemic-v.0.1]), it was to assess the Lumipulse assay’s results with respect to the results of SARS-CoV-2 subgenomic RNA assay (14), which was performed on the samples concomitantly tested for SARS-CoV-2 antigen. Thus, among 194 (SARS-CoV-2 genomic RNA) RT-PCR positive samples, 101 (52.1%) samples had positive results for subgenomic RNA (and antigen), with 82 (94.3%) of 87 antigen-positive samples having RT-PCR Ct values below 18–25 (Tables 2 and 3). Importantly, 54 of 93 samples negative at the subgenomic RNA assay but positive at the Lumipulse antigen assay included samples from patients who were tested 6.1 ± 7.0 days after COVID-19 diagnosis (Table 3), suggesting that SARS-CoV-2 antigen may be longer detected than SARS-CoV-2 subgenomic RNA. Consistent with these findings, 101 aforementioned samples were from patients who were tested 1.6 ± 3.3 days after COVID-19 diagnosis (Table 3), suggesting that SARS-CoV-2 antigen may indicate active infection. It should be recalled that SARS-CoV-2 virions do not contain subgenomic RNA—i.e., RNA replicative forms thought to encode the structural spike (S), E, membrane (M), or N virus proteins—whereas subgenomic RNA is part of cellular membrane vesicles and thereby relatively stable (15). Thus, antigen and subgenomic RNA represent two virus biology entities worthy of investigation in clinical samples (7, 18, 19), especially in situations of prolonged (genomic RNA) RT-PCR positivity implying infectious virus shedding (15).

In an attempt to appraise fully Lumipulse assay’s performance, we analyzed 194 RT-PCR positive samples stratified by groups of testing (Table 2). This stratification allowed us to assess the accuracy of the Lumipulse assay to determine if a person presenting in the (primary or secondary care) hospital or the community has SARS-CoV-2 infection. Therefore, we included 194 adults suspected of (*n* = 69) or screened for (*n* = 53) SARS-CoV-2 infection or monitored for confirmed COVID-19 (*n* = 72) in diagnostic, screening, or monitoring groups, respectively. We found that the Lumipulse assay worked well, and almost equally, in all three testing groups, with 60 (diagnostic group), 43 (screening group), or 52 (monitoring group) samples being positive. Expectedly, the subgenomic RNA assay yielded positive results in 44 (diagnostic group), 33 (screening group), or 24 (monitoring group) samples. Of note, lowest sample positivity rates were seen in the monitoring group with both Lumipulse (52/72 samples, 72.2%) and subgenomic RNA (24/72 samples, 33.3%) assays. These findings concur with the idea that SARS-CoV-2 antigen or, particularly, subgenomic RNA results are likely to be less positive in monitoring scenarios where positive results for genomic RNA are, instead, indicative of prolonged SARS-CoV-2 shedding (15). Accordingly, in our monitoring group, the time from COVID-19 diagnosis to testing was longer (up to 32 days) than in diagnostic or screening groups (0 days).

To the best of our knowledge, this is to date the largest clinical study evaluating the Lumipulse assay. Compared to previous studies (6, 7), our set of tested nasopharyngeal swab samples is not only wider but also uncharted—we included 594 individuals’ samples from testing scenarios with different pretest probability that, in turn, reflected different clinical situations. Nonetheless, our findings agreed with those by Hirotsu et al. (6) showing that the SARS-CoV-2 antigen levels declined in consecutively collected samples of seven patients from the time of their hospital admission to discharge. Therefore, the finding that antigen positivity rates varied according to whether samples were in a diagnostic/screening rather than in a monitoring scenario reinforces the hypothesis raised by Hirotsu et al. that antigen testing could be also useful to identify patients in the early or late phase of SARS-CoV-2 infection (6). In an evaluations’ review of five antigen tests (four commercial and one in-house) by Dinnes et al. (17), average sensitivity was 56.2% (95% CI, 29.5 to 79.8%) and average specificity was 99.5% (95% CI 98.1 to 99.9%) based on five studies with 943 samples (596 were confirmed SARS-CoV-2 samples). However, the sensitivity varied considerably across studies (from 0% to 94%), causing uncertainty about how useful antigen tests are in clinical practice (17). To enhance the applicability of our Lumipulse assay’s results, we determined the assay’s analytical sensitivity (Fig. S1) before sample testing to ensure that the assay’s LOD was equivalent to 10^4^ viral genomic copies/ml, which is the desirable limit acknowledged to date (https://www.who.int/publications/m/item/covid-19-target-product-profiles-for-priority-diagnostics-to-support-response-to-the-covid-19-pandemic-v.0.1). Furthermore, we limited oversampling of samples with laboratory-confirmed SARS-CoV-2 infection, which accounted for the high risk of bias affecting patient selection in many published studies (17). Stratifying our study participants by days from the symptom onset (5) was impracticable for us. However, we compensated for this limitation by including testing groups that were comparable for size (∼60 RT-PCR positive samples per group), and we assumed that RT-PCR negative samples were almost equally distributed across testing groups.

To summarize, our results show that Lumipulse assay’s performance was satisfactory, confirming the current view about antigen-based laboratory testing for SARS-CoV-2 detection. In particular, the Lumipulse assay was highly sensitive to detect SARS-CoV-2 antigen in samples with low RT-PCR Ct values (<25) by overall or different testing scenarios. While Ct values >25 might not correspond to situations with active SARS-CoV-2 infection and/or infectivity, a strategy of repeated testing can maximize the Lumipulse assay’s performance and thereby reduce consequences of false-negative results.

## Data Availability

All the data presented in the study are available on request.

## ACKNOWLEDGEMENTS

The authors are grateful to the Reale Group and the Fondazione Valentino Garavani & Giancarlo Giammetti for providing financial support to the COVID-19 Research at the FPG (<u>F</u>ondazione <u>P</u>oliclinico Universitario A. <u>G</u>emelli IRCCS) of Rome (Italy), or with the Italian Health Ministry for providing funds to the INMI (<u>I</u>stituto <u>N</u>azionale per le <u>M</u>alattie <u>I</u>nfettive Lazzaro Spallanzani IRCCS) of Rome (Italy).

We thank Franziska Lohmeyer for English revision of the manuscript.

---

**FIG S1** Probit analysis to calculate the limit of detection (LOD) of the Lumipulse G SARS-CoV-2 Ag assay. In this analysis, input was the numbers of samples with positive detection, which were obtained with Vero E6 cell-cultured SARS-CoV-2 (INMI-1 strain) tested in replicates at a concentration range of 1.0 × 10^5^ 50% tissue culture infective dose (TCID_50_)/ml (4.0 × 10^8^ RNA copies/ml) to 1.0 TCID_50_/ml (4.0 × 10^3^ RNA copies/ml). The output was the Lumipulse assay’s LOD, which was equivalent to a value of 0.47 log_10_ TCID_50_/ml (otherwise expressed as 2.95 TCID_50_/ml) corresponding to 4.07 log_10_ RNA copies/ml (otherwise expressed as 1.2 × 10^4^ copies/ml)

## REFERENCES

1. Brooks ZC, Das S. 2020. COVID-19 testing. Am J Clin Pathol 154:575–584. https://doi.org/10.1093/ajcp/aqaa141.

2. Cheng MP, Papenburg J, Desjardins M, Kanjilal S, Quach C, Libman M, Dittrich S, Yansouni CP. 2020. Diagnostic testing for severe acute respiratory syndrome–related coronavirus-2: a narrative review. Ann Intern Med M20–1301. https://doi.org/10.7326/M20-1301.

3. Mak GC, Lau SS, Wong KK, Chow NL, Lau CS, Lam ET, Chan RC, Tsang DN. 2020. Analytical sensitivity and clinical sensitivity of the three rapid antigen detection kits for detection of SARS-CoV-2 virus. J Clin Virol 133:104684. https://doi.org/10.1016/j.jcv.2020.104684.

4. Ogawa T, Fukumori T, Nishihara Y, Sekine T, Okuda N, Nishimura T, Fujikura H, Hirai N, Imakita N, Kasahara K. 2020. Another false-positive problem for a SARS-CoV-2 antigen test in Japan. J Clin Virol 131:104612. https://doi.org/10.1016/j.jcv.2020.104612.

5. European Centre for Disease Prevention and Control (ECDC). Options for the use of rapid antigen tests for COVID-19 in the EU/EEA and the UK. 19 November 2020. ECDC: Stockholm; 2020.

6. Hirotsu Y, Maejima M, Shibusawa M, Nagakubo Y, Hosaka K, Amemiya K, Sueki H, Hayakawa M, Mochizuki H, Tsutsui T, Kakizaki Y, Miyashita Y, Yagi S, Kojima S, Omata M. 2020. Comparison of automated SARS-CoV-2 antigen test for COVID-19 infection with quantitative RT-PCR using 313 nasopharyngeal swabs, including from seven serially followed patients. Int J Infect Dis 99:397–402. https://doi.org/10.1016/j.ijid.2020.08.029.

7. Hirotsu Y, Maejima M, Shibusawa M, Amemiya K, Nagakubo Y, Hosaka K, Sueki H, Hayakawa M, Mochizuki H, Tsutsui T, Kakizaki Y, Miyashita Y, Omata M. 2020. Analysis of a persistent viral shedding patient infected with SARS-CoV-2 by RT-qPCR, FilmArray Respiratory Panel v2.1, and antigen detection. J Infect Chemother Oct 29:S1341-321X(20)30396-2. https://doi.org/10.1016/j.jiac.2020.10.026. Epub ahead of print.

8. Liotti FM, Menchinelli G, Marchetti S, Morandotti GA, Sanguinetti M, Posteraro B, Cattani P. 2020. Evaluation of three commercial assays for SARS-CoV-2 molecular detection in upper respiratory tract samples. Eur J Clin Microbiol Infect Dis Sep 4:1–9. https://doi.org/10.1007/s10096-020-04025-0. Epub ahead of print.

9. Liotti FM, Menchinelli G, Lalle E, Palucci I, Marchetti S, Colavita F, La Sorda M, Sberna G, Bordi L, Sanguinetti M, Cattani P, Capobianchi MR, Posteraro B. 2020. Performance of a novel diagnostic assay for rapid SARS-CoV-2 antigen detection in nasopharynx samples. Clin Microbiol Infect Sep 23:S1198-743X(20)30583-8. https://doi.org/10.1016/j.cmi.2020.09.030. Epub ahead of print.

10. Poljak M, Korva M, Knap Gašper N, Fujs Komloš K, Sagadin M, Uršič T, Avšič Županc T, Petrovec M. 2020. Clinical evaluation of the Cobas SARS-CoV-2 test and a diagnostic platform switch during 48 hours in the midst of the COVID-19 pandemic. J Clin Microbiol 58:e00599–20. https://doi.org/10.1128/JCM.00599-20.

11. Procop GW, Brock JE, Reineks EZ, Shrestha NK, Demkowicz R, Cook E, Ababneh E, Harrington SM. A 2021. Comparison of five SARS-CoV-2 molecular assays with clinical correlations. Am J Clin Pathol 155:69–78. https://doi.org/10.1093/ajcp/aqaa181.

12. Liotti FM, Menchinelli G, Marchetti S, Morandotti GA, Sanguinetti M, Posteraro B, Cattani P. 2020. Evaluating the newly developed BioFire COVID-19 test for SARS-CoV-2 molecular detection. Clin Microbiol Infect 26:1699–1700. https://doi.org/10.1016/j.cmi.2020.07.026.

13. Liotti FM, Menchinelli G, Marchetti S, Posteraro B, Landi F, Sanguinetti M, Cattani P. 2020. Assessment of SARS-CoV-2 RNA test results among patients who recovered from COVID-19 with prior negative results. JAMA Intern Med Nov 12:e207570. https://doi.org/10.1001/jamainternmed.2020.7570. Epub ahead of print.

14. Wölfel R, Corman VM, Guggemos W, Seilmaier M, Zange S, Müller MA, Niemeyer D, Jones TC, Vollmar P, Rothe C, Hoelscher M, Bleicker T, Brünink S, Schneider J, Ehmann R, Zwirglmaier K, Drosten C, Wendtner C. 2020. Virological assessment of hospitalized patients with COVID-2019. Nature 581:465–469. https://doi.org/10.1038/s41586-020-2196-x.

15. Alexandersen S, Chamings A, Bhatta TR. 2020. SARS-CoV-2 genomic and subgenomic RNAs in diagnostic samples are not an indicator of active replication. Nat Commun 2020 11:6059. https://doi.org/10.1038/s41467-020-19883-7.

16. Clerc O, Greub G. 2010. Routine use of point-of-care tests: usefulness and application in clinical microbiology. Clin Microbiol Infect 2010 16:1054–1061. https://doi.org/10.1111/j.1469-0691.2010.03281.x.

17. Dinnes J, Deeks JJ, Adriano A, Berhane S, Davenport C, Dittrich S, Emperador D, Takwoingi Y, Cunningham J, Beese S, Dretzke J, Ferrante di Ruffano L, Harris IM, Price MJ, Taylor-Phillips S, Hooft L, Leeflang MM, Spijker R, Van den Bruel A; Cochrane COVID-19 Diagnostic Test Accuracy Group. 2020. Rapid, point-of-care antigen and molecular-based tests for diagnosis of SARS-CoV-2 infection. Cochrane Database Syst Rev 8:CD013705. https://doi.org/10.1002/14651858.CD013705.

18. Rodríguez-Grande C, Adán-Jiménez J, Catalán P, Alcalá L, Estévez A, Muñoz P,Pérez-Lago L, de Viedma DG; Gregorio Marañón Microbiology-ID COVID 19 Study Group. 2020. Inference of active viral replication in cases with sustained positive RT-PCRs for SARS-CoV-2. J Clin Microbiol Nov 25:JCM.02277-20. https://doi.org/10.1128/JCM.02277-20. Epub ahead of print.

19. Avanzato VA, Matson MJ, Seifert SN, Pryce R, Williamson BN, Anzick SL, Barbian K, Judson SD, Fischer ER, Martens C, Bowden TA, de Wit E, Riedo FX, Munster VJ. 2020. Case Study: Prolonged Infectious SARS-CoV-2 Shedding from an Asymptomatic Immunocompromised Individual with Cancer. Cell 183:1901-1912.e9. doi: 10.1016/j.cell.2020.10.049.

